# Advancing Efficacy Prediction for EHR-based Emulated Trials in Repurposing Heart Failure Therapies

**DOI:** 10.1101/2023.05.25.23290531

**Authors:** Nansu Zong, Shaika Chowdhury, Shibo Zhou, Sivaraman Rajaganapathy, Yue Yu, Liewei Wang, Qiying Dai, Pengyang Li, Xiaoke Liu, Suzette J. Bielinski, Jun Chen, Yongbin Chen, James R. Cerhan

## Abstract

**Introduction:** The High mortality rates associated with heart failure (HF) have propelled the strategy of drug repurposing, which seeks new therapeutic uses for existing, approved drugs to enhance the management of HF symptoms effectively. An emerging trend focuses on utilizing real-world data, like EHR, to mimic randomized controlled trials (RCTs) for evaluating treatment outcomes through what are known as emulated trials (ET). Nonetheless, the intricacies inherent in EHR data—comprising detailed patient histories in databases, the omission of certain biomarkers or specific diagnostic tests, and partial records of symptoms— introduce notable discrepancies between EHR data and the stringent standards of RCTs. This gap poses a substantial challenge in conducting an ET to accurately predict treatment efficacy.

**Objective:** The objective of this research is to predict the efficacy of drugs repurposed for HF in randomized trials by leveraging EHR in ET.

**Methods:** We proposed an ET framework to predict drug efficacy, integrating target prediction based on biomedical databases with statistical analysis using EHR data. Specifically, we developed a novel target prediction model that learns low-dimensional representations of drug molecules, protein sequences, and diverse biomedical associations from a knowledge graph. Additionally, we crafted strategies to improve the prediction by considering the interactions between HF drugs and biological factors in the context of HF prognostic markers.

**Results:** Our validation of the drug-target prediction model against the BETA benchmark demonstrated superior performance, with an average AUCROC of 97.7%, PRAUC of 97.4%, F1 score of 93.1%, and a General Score of 96.1%, surpassing existing baseline algorithms. Further analysis of our ET framework on identifying 17 repurposed drugs—derived from 266 phase 3 HF RCTs—using data from 59,000 patients at the Mayo Clinic highlighted the framework’s remarkable predictive accuracy. This analysis took into account various factors such as biological variables (e.g., gender, age, ethnicity), HF medications (e.g., ACE inhibitors, Beta-blockers, ARBs, Loop Diuretics), types of HF (HFpEF and HFrEF), confounders, and prognostic markers (e.g., NT-proBNP, BUN, creatinine, and hemoglobin). The ET framework significantly improved the accuracy compared to the baseline efficacy analysis that utilized EHR data. Notably, the best results were improved in AUC-ROC from 75.71% to 93.57% and in PRAUC from 78.66% to 90.34%, compared to the baseline models.

**Conclusion:** Our study presents an ET framework that significantly enhances drug efficacy emulation by integrating EHR-based analysis with target prediction. We demonstrated substantial success in predicting the efficacy of 17 HF drugs repurposed for phase 3 RCTs, showcasing the framework’s potential in advancing HF treatment strategies.

## 1. Introduction

Heart failure (HF) is a critical medical condition with high rates of hospitalization and mortality^1^, where the heart fails to adequately pump blood to meet the body’s needs^2^. There is an urgent need for improved therapeutic approaches to effectively manage HF symptoms, enhance the quality of life, and increase survival rates. Drug repurposing is an innovative approach to drug discovery that involves finding new therapeutic uses for existing drugs^3^. Since the safety profiles of the drugs (e.g., pharmacokinetics/pharmacodynamics) for repurposing are already established through rigorous tests in the early phases^4, 5^, evaluating the potential efficacy of the repurposed drug candidate is critical to streamline the process and efficiently allocate resources for the large-scale trials like phase 3 clinical trials, which can be challenging (e.g., 20% failure rate for HF^6^) and costly due to multiple factors ^7^.

A growing trend involves leveraging real-world data, such as Electronic Health Records (EHR), to simulate randomized trials (RCTs) for assessing treatment outcomes, referred to as emulated trials (ET) ^8, 9, 10, 11, 12, 13, 14, 15, 16, 17^. Contrary to the controlled settings of RCTs, ETs utilize observational data from everyday clinical practices. This data encompasses the complexities of patient histories stored in databases, the absence of certain biomarkers or specific diagnostic images, and incomplete records of symptoms^15^. Although ETs are theoretically expected to adhere to the same eligibility criteria as RCTs, within the ‘target trial framework,’^10^ to select appropriate patient groups and extract relevant data from EHR for efficacy evaluation, the discrepancies^14, 15^ between RCT standards and EHR data present a significant challenge in executing an ET effectively for predicting treatment effectiveness^18, 19, 20, 21, 22, 23, 24^. Although the existing cutting-edge ET methods^25, 26^ have introduced AI to balance confounders and simplify criteria for patient selection, their primary focus has not been on improving the accuracy of ET predictions, and their performance in efficacy prediction is not been tested.

This study introduces a novel framework designed to predict the efficacy of repurposed drugs based on EHR. Specifically, the framework improves ET by integrating a proposed target prediction algorithm that utilizes drug chemical compositions, protein sequences, and heterogeneous biomedical knowledge bases. We evaluated the target prediction algorithm using a significant drug-target prediction benchmark known as BETA^27^. For evaluating the proposed ET framework, we manually identified 17 repurposed drugs for HF from 266 phase 3 RCTs, which included 7 with a positive effect and 10 without. Our in-depth analysis covered multiple HF-related clinical indicators (e.g., N-terminal pro-b-type natriuretic peptide, blood urea nitrogen, Creatinine, and hemoglobin), patient cohorts (e.g., Heart failure with preserved ejection fraction and Heart failure with reduced ejection fraction), HF medications (e.g., angiotensin-converting-enzyme inhibitors, beta-blockers, angiotensin II receptor blockers, and loop diuretics), and biological factors (e.g., sex). The ET framework significantly improved the predictive accuracy over the baseline efficacy analysis using EHR data, as evidenced in AUCROC (P-value = 2.2e-16) and PRAUC (P-value = 2.2e-16). Notably, the best results were increased from 75.71% to 93.57% for AUC-ROC and from 78.66% to 90.34% for PRAUC.

## 2. Results

### 2.1. Proposed framework-integration EHR-based efficacy analysis with target prediction

Our proposed an ET framework that integrates target prediction with EHR-based drug-efficacy analysis. The process comprises four key steps (refer to **Figure 1 (a)** for the study design): 1) identifying genes associated with the disease (i.e., HF), 2) predicting drug targets based on HF-related genes, 3) extracting the study cohort from EHR, and 4) efficacy analysis.

**Figure 1.**
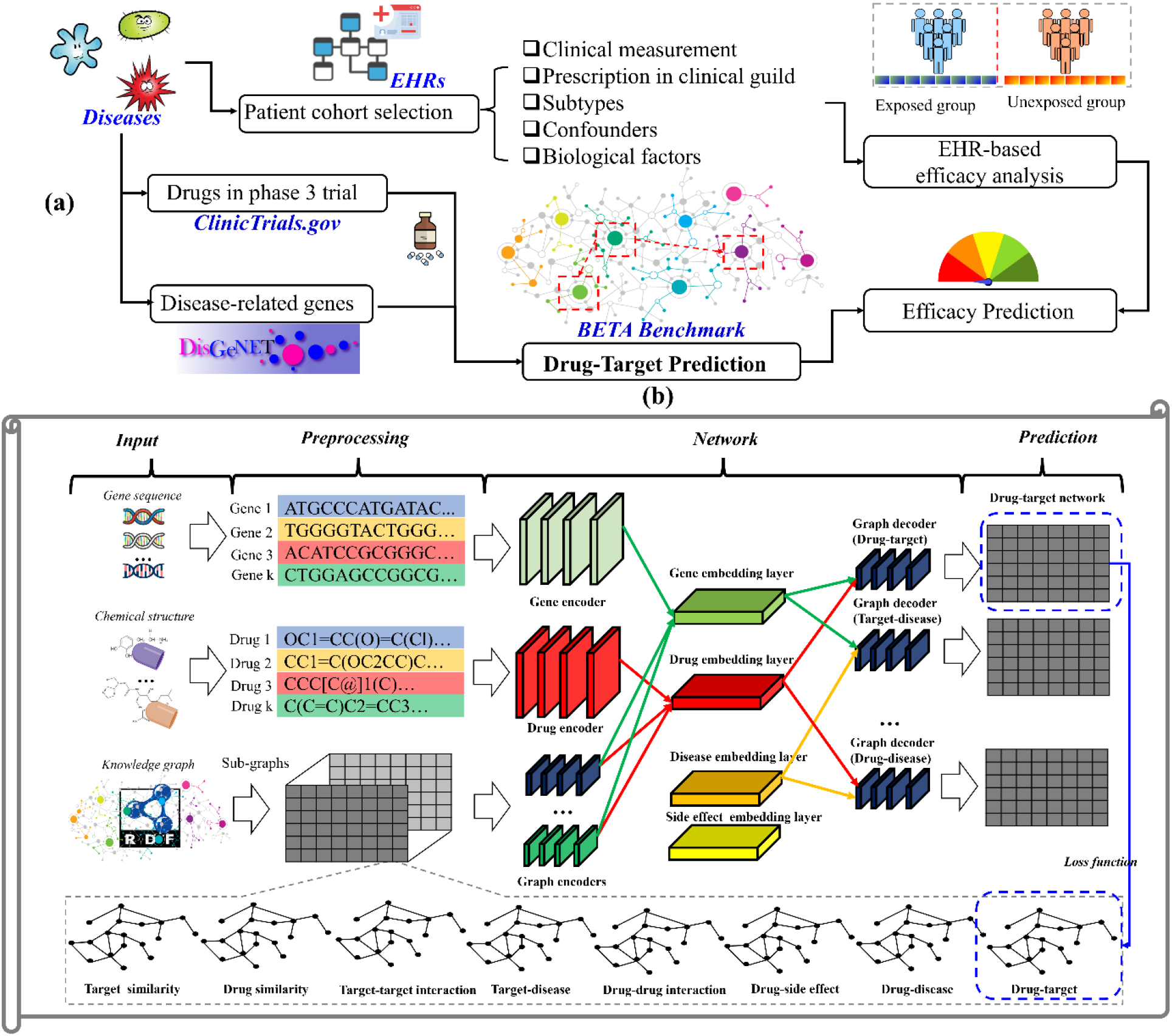
An overview of the proposed framework. **(a)** shows the general pipeline of the framework, which starts with a search for the relevant clinical trial drugs on ClinicTrials.gov and the identification of genes using the DisGeNET^28^ database. Using a list of repurposed trial drugs and the genes, the proposed target prediction model is used to predict the probabilities of drug-gene associations. The efficacy of the repurposed drug is then analyzed based on EHR data, where the exposed group and unexposed groups are compared against a list of clinical tests related to HF. The overall prediction results are obtained by combining the results from target prediction and EHR-based analysis. **(b)** provides more details about the developed target prediction model, which employs drug chemical structure, protein sequence, and knowledge graph to obtain embedding layers for each biomedical entity. The cost function is calculated by minimizing the weights of the re-constructed sub-knowledge graphs and original graphs. The predicted results are extracted from the re-constructed drug-target graph.

In the initial phases, we developed an innovative algorithm for predicting drug targets. This algorithm is designed to assimilate information regarding the chemical structure of drugs, the sequence of proteins, and biomedical relationships, with the aim of reconstructing the original knowledge graph. This reconstruction facilitates the prediction of new potential associations between drugs and their targets. In the subsequent stages, our research made use of the Mayo Clinic database, which comprises a wide range of variables. These include three biological factors—age, sex, and ethnicity—as well as various HF medications such as angiotensin-converting-enzyme inhibitors (ACEI), beta-blockers (BB), angiotensin II receptor blockers (ARB), and loop diuretics (LD). We also explored HF subtypes, such as HF with reduced ejection fraction (HFrEF) and HF with preserved ejection fraction (HFpEF), and included BASELINE cohorts, which are specific cohorts (e.g., HFrEF) identified by the inclusion criteria in RCTs. The Mayo Clinic patient database encompasses a total of 59,102 HF patients. For statistical details, please refer to **Table 1**. The EHR data will be employed to improve the prediction of efficacy using the target prediction algorithm. For a comprehensive explanation of these methods, please see **Section 4**.

**Table 1.** Statistics of the HF patients from the Mayo Clinic EHR.

| **Variables** | **# patient** | **Variables** | **# patient** |
| --- | --- | --- | --- |
| **Age ± SD** | 53.0 ± 30.9 | **Gender** | |
| **Ethnicity** |  | Female | 17,931 |
| Non-Hispanic White | 37,406 | Male | 24,005 |
| Others | 4,531 | **Prognostic Markers** |  |
| Unknown | 17,165 | NT-proBNP | 25,567 |
| **Baseline HF drugs** |  | Creatinine | 36,731 |
| ACEI | 26,306 | BUN | 41,598 |
| BB | 45,404 | Hemoglobin | 41,626 |
| ARB | 17,240 | **Repurposed drugs** |  |
| LD | 52,889 | Albuterol | 32,697 |
| **HF subtypes** |  | Amiodarone | 16,519 |
| HFpEF | 33,259 | Aspirin | 45,911 |
| HFrEF | 28,123 | Cholecalciferol | 12,675 |
| **Confounders** |  | Dexamethasone | 30,825 |
| Arrhythmias (including baseline variations, Atrial fibrillation, Flutter) | 29,268 | Macitentan | 203 |
| Cardiac Arrest (including history of) | 1,870 | Metformin | 5,182 |
| Myocardial infarction (past, prior) | 12,935 | Methotrexate | 698 |
| Hypertension (including long-standing) | 33,602 | Mirabegron | 405 |
| Pulmonary hypertension | 2,619 | Prednisone | 16,401 |
| Ischemic heart disease | 27,814 | Ranolazine | 811 |
| Hypertensive heart and Renal disease | 3,671 | Rosuvastatin | 7,384 |
| Medications affecting heart health | 777 | Sertraline | 6,306 |
| Congestive heart failure, unspecified | 30,819 | Sildenafil | 800 |
| Renal Failure (including end-stage, acute, and chronic) | 22,137 | Simvastatin | 9,819 |
| Diabetes Mellitus (type 1 and type 2) | 17,572 | Thiamine | 5,860 |
| **# HF in total** | 59,102 | Warfarin | 21,291 |

### 2.2. Target prediction model and the evaluation with BETA benchmark

Our proposed target prediction model was put to the test using the BETA benchmark, which comprises seven tests and 344 tasks designed to evaluate drug-target prediction models. These tests evaluate the models from various perspectives, including general screening, target and drug screening based on categories, searching for specific drugs and targets, and drug repurposing for specific diseases. To compare our model’s performance, we used six state-of-the-art predictive models (DTINet^29^, bioLNE^30^, NeoDTI^31^, DeepPurpose^32^, DeepDTA^33^, GraphDTA^34^) as baseline methods (See **Figure 2**). The proposed method performed exceptionally well with an average AUCROC of 97.7%, PRAUC of 97.4%, F1 score of 93.1%, and General Score of 96.1%, outperforming the best-performing baseline methods, DeepPurpose (average AUCROC: 88.0%, PRAUC: 84.4%, F1: 79.5%, General Score: 84.0%), and NeoDTI (average AUCROC: 86.5%, PRAUC: 83.3%, F1: 74.2%, General Score: 81.3%). Our proposed method was outstanding, with 266 out of 404 tasks (65.8%) outperforming other methods across all seven tests. It’s important to note that for test 4, we counted the average precision for the top 10, 20, 50, and 100, which made 80 tasks counted rather than 20 tasks counted in the BETA. For each test, our proposed method achieved excellent results with 10/10 (100.0%), 61/90 (67.8%), 66/72(91.7%), 62/72(86.1%), 42/80 (52.5%), 17/40 (42.5%), 8/40 (20.0%). Please refer to the **Supplementary Figures 1-11** for more detailed results.

**Figure 2.**
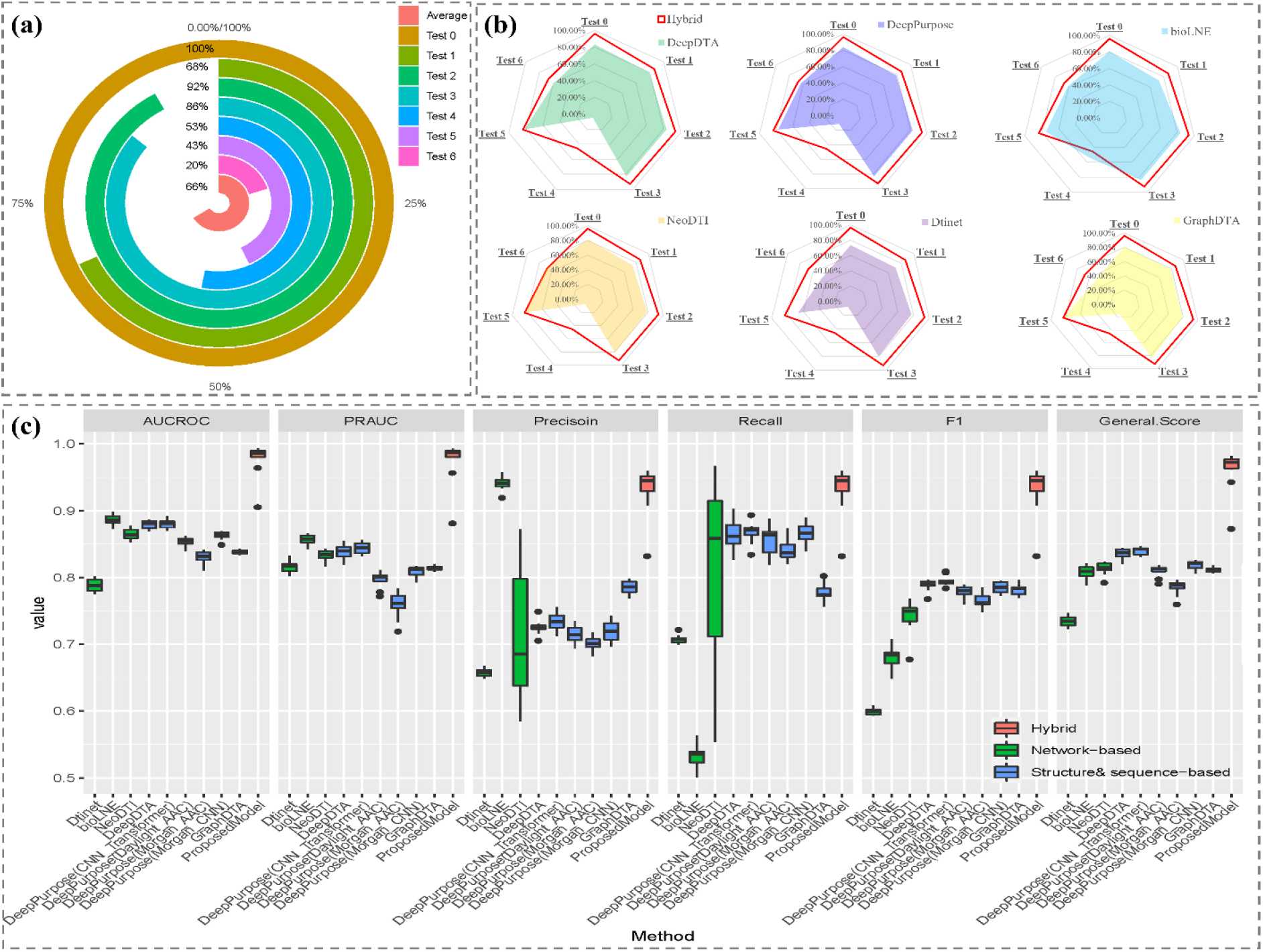
Performance of the proposed drug-target prediction method compared to other baseline methods in the evaluation based on the BETA benchmark. **(a)** the identification of the best-performing method among all comparison methods for each test in the BETA evaluation tasks, **(b)** a 1-to-1 comparison between the proposed method (represented by the red line) and the baseline method (represented by the colored area) in all the tests on the spider chart, and **(c)** a breakdown of the detailed results of the overall test in BETA evaluation tasks (i.e., Test 0). Overall, the proposed method outperforms all other methods across all evaluation measurements.

### 2.3. EHR-based efficacy analysis

To assess the efficacy of 17 drugs repurposed for phase 3 HF clinical trials, where 7 demonstrated positive outcomes and 10 showed no effect, we performed chi-square tests across various combinations of cohorts (namely BASELINE, ALL, HFpEF, and HFrEF) and prognostic markers (NT-proBNP, BUN, Creatinine, Hemoglobin). As illustrated in **Figure 3**, the combination of BASELINE cohorts and NT-proBNP marker emerged as the most predictive of drug efficacy (with AUC-ROC=75.71% and PRAUC=78.66%) according to the chi-square tests. Although most drugs that showed no effect were accurately predicted as such, a large number of drugs that had positive effects were incorrectly predicted, indicating a high rate of false positives for the efficacy prediction only using EHR. For the whole combinations for cohorts and prognostic markers, please refer to **Supplementary Figures13**.

**Figure 3.**
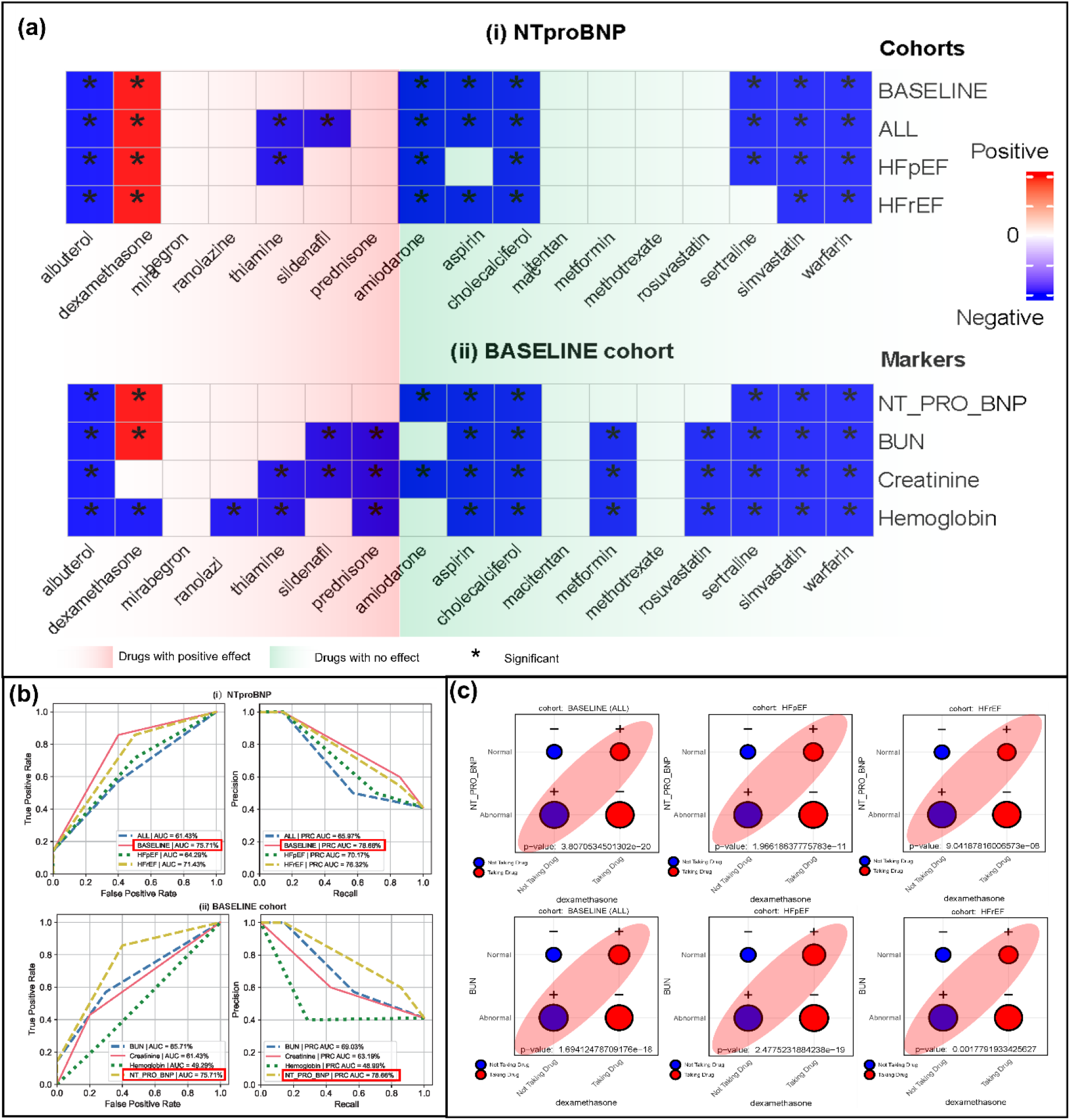
Evaluation of treatment efficacy from EHR Data. (a) presents all analyses conducted for (i) four distinct groups (i.e., Baseline group, all HF patients, HFpEF, and HFrEF) using NT-proBNP, and (ii) four prognostic markers (specifically, NT-proBNP, BUN, Creatinine, Hemoglobin) for the BASELINE patient group. (b) Analysis outcomes in terms of AUC-ROC and PRAUC metrics for the evaluations in (i) and (ii). (c) The detected positive association between the prognostic marker levels and the use of the repurposed drugs under study, as determined through the Chi-square test. The ‘+’ and ‘-’ symbols indicate that the number of observed patients is greater than or less than expected, respectively. The areas shaded in red demonstrate a positive correlation between the administration of the drug and an elevated-normal level of a specific prognostic marker.

### 2.4. Improving efficacy prediction with integration of target prediction

The predictions of 17 drugs tested against 25 HF-related genes related to HF are displayed in **Figure 4 (a)**. Dexamethasone, thiamine B1, ranolazine and are the top drugs predicted to have the most HF-related genes, with 24/25, 20/25, and 15/25 genes, respectively. On the other hand, Simvastatin, Warfarin, and Rosuvastatin have the least number of HF-related genes, with only 2/25, 2/25, 2/25, and 1/25 genes, respectively. It is worth noting that all these drugs are used for treating or managing medical conditions related to the cardiovascular system. Specifically, Rosuvastatin and Simvastatin work by reducing cholesterol production and increasing clearance of LDL-cholesterol from the bloodstream by inhibiting HMG-CoA reductase, an enzyme involved in cholesterol synthesis. Regarding genes, NOS3, ADRB1, and VEGFA have the most HF-repurposed drugs, with 14/24, 12/24, and 11/24 drugs, respectively. Meanwhile, PTH, RAC 1, and GRK2 are the genes with the least number of repurposed drugs, with only 1/24 drugs for each. We show the performance of efficacy prediction based on target prediction in **Supplementary Figures12**. We also combined target prediction outcomes with chi-square tests for better efficacy forecasts. **Figures 4(b) and (c)** highlight the benefits of incorporating target prediction into efficacy forecasts, notably a decrease in false positives, as drugs showing positive effects are often associated with a higher count of positive targets. In particular, for NT-proBNP within the BASELINE cohort, there’s an improvement from an AUC-ROC of 75.71% and a PRAUC of 78.66% to an AUC-ROC of 85.71% and a PRAUC of 87.28%. The integration of target prediction significantly improves EHR-based analysis, as demonstrated in Section 2.3. This improvement is evident in a single-tailed T-test for AUCROC (P-value = 4.205e-12) and PRAUC (P-value = 2.743e-06). For the whole combinations for cohorts and prognostic markers, please refer to **Supplementary Figures14**.

**Figure 4.**
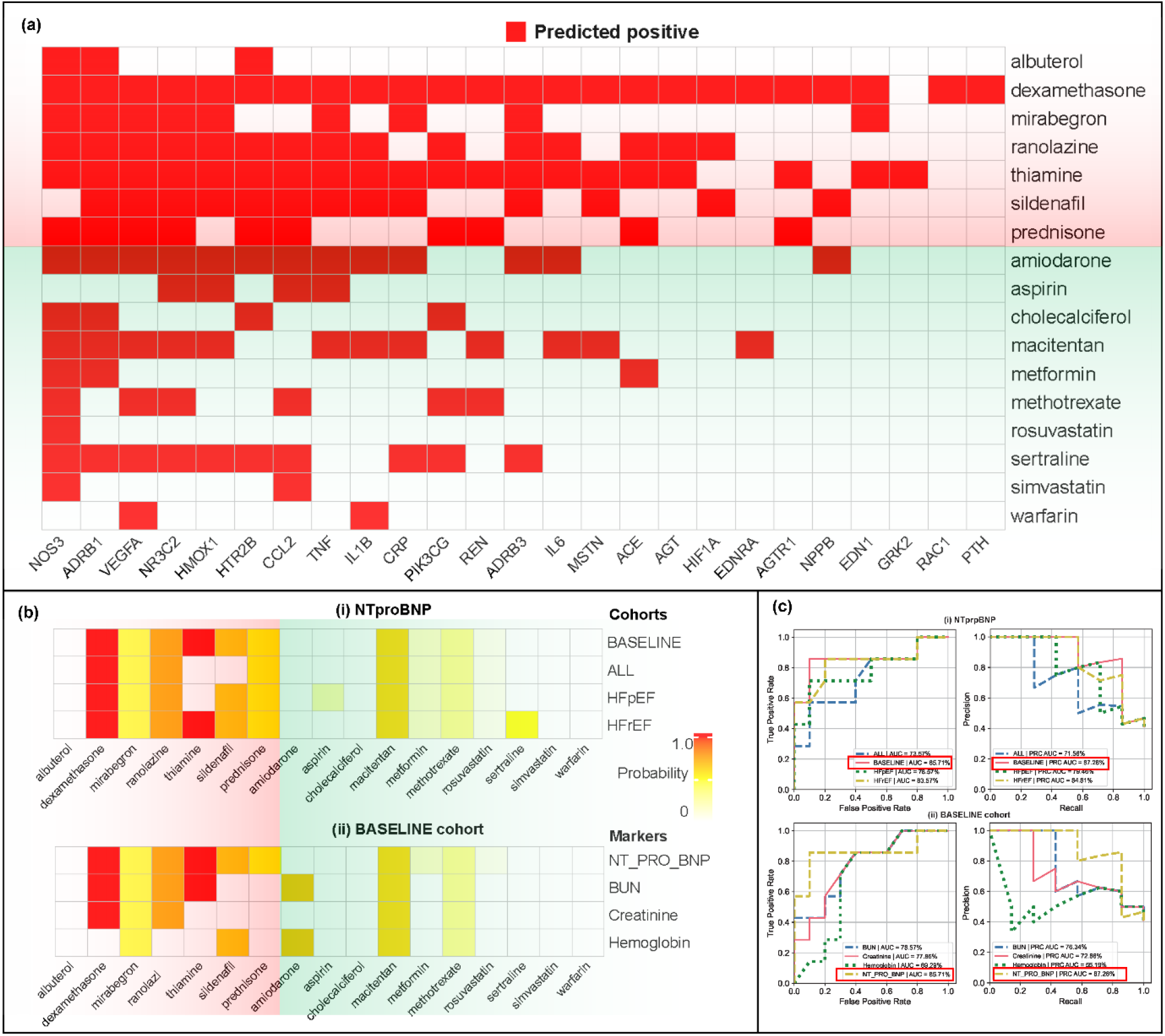
Efficacy predictions based on the integration of target predictions and EHR-based analysis. (a) shows the target prediction for 17 drugs based on the proposed algorithm. (b) presents a chi-square analysis for four distinct groups using NT-proBNP, and (ii) four prognostic markers for the BASELINE patient group. (c) presents the outcomes in terms of AUC-ROC and PRAUC metrics for the evaluations in (i) and (ii).

### 2.5. Improvement by considering Interaction with HF drugs and Sex

To refine our predictive model by accounting for variable interactions, we incorporated the coefficients derived from the binomial regression model into our target prediction. This approach was employed particularly when a significant correlation (i.e., P-value<0.5) was observed between the variation in biomarkers and the intake of drugs, thereby enabling us to more accurately generate efficacy predictions. For each prognostic marker, we factored in the interactions between HF medications and gender, which showed all the increases and decreases in outcomes in **Figure 5 (a)**. Among the four markers, aside from NT-proBNP, the other three indicators (BUN, Creatinine, and Hemoglobin) demonstrated enhanced predictive accuracy when accounting for interactions. No significant advantages were noted for NT-proBNP upon including interactions. Notably, BUN and Hemoglobin showed considerable improvements, with BUN and Hemoglobin accounting for 6 and 11 of the top 23 results, respectively. Further detailed analysis presented in **Figure 5 (b)** highlights that the best interaction combinations elevated the AUCROC to 93.57% and PRAUC to 90.34%, up from 85.71% and 87.28%. This improvement of including interactions in efficacy prediction is evident in a single-tailed T-test for AUCROC (P-value = 8.338e-08) and PRAUC (P-value = 2.004e-15). For the improvement evaluated by PRAUC, please refer to **Supplementary Figure 15**.

**Figure 5.**
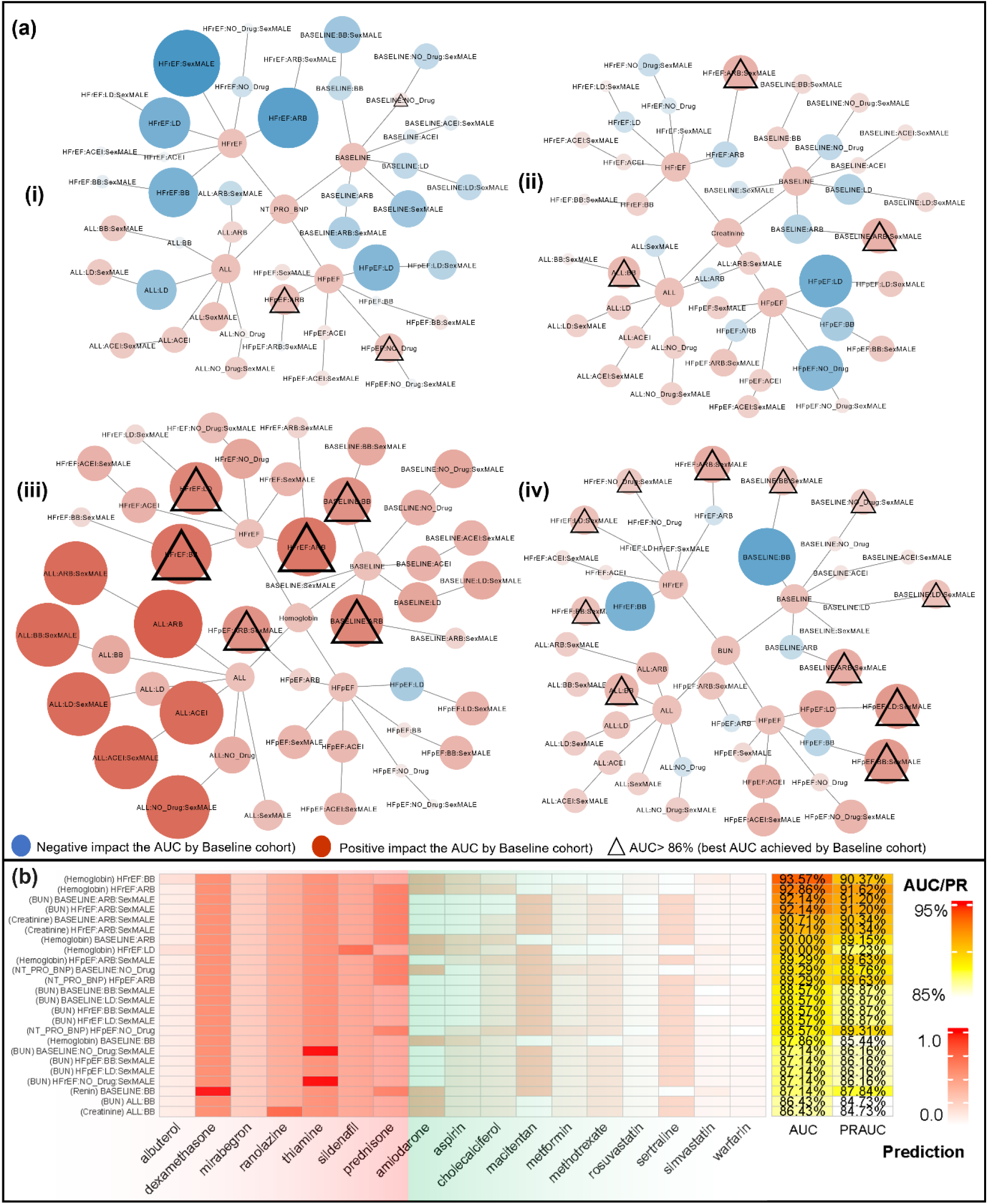
Improvement of including the interactions of HF Drugs and Gender in EHR-based analysis. (a) displays improved predictions through various interaction combinations for our prognostic markers, (i) NT-proBNP Creatinine and (iii) Hemoglobin, and (iv) BUN. Red nodes symbolize improvement, while blue nodes depict a decrease in AUCROC performance. The triangle marks the highest AUCROC improvement. (b) details the prediction for 17 drugs in the most effectively performing combinations (as indicated by triangles in (a)) along with the respective AUCROC and PRAUC metrics.

**Figure 6.**
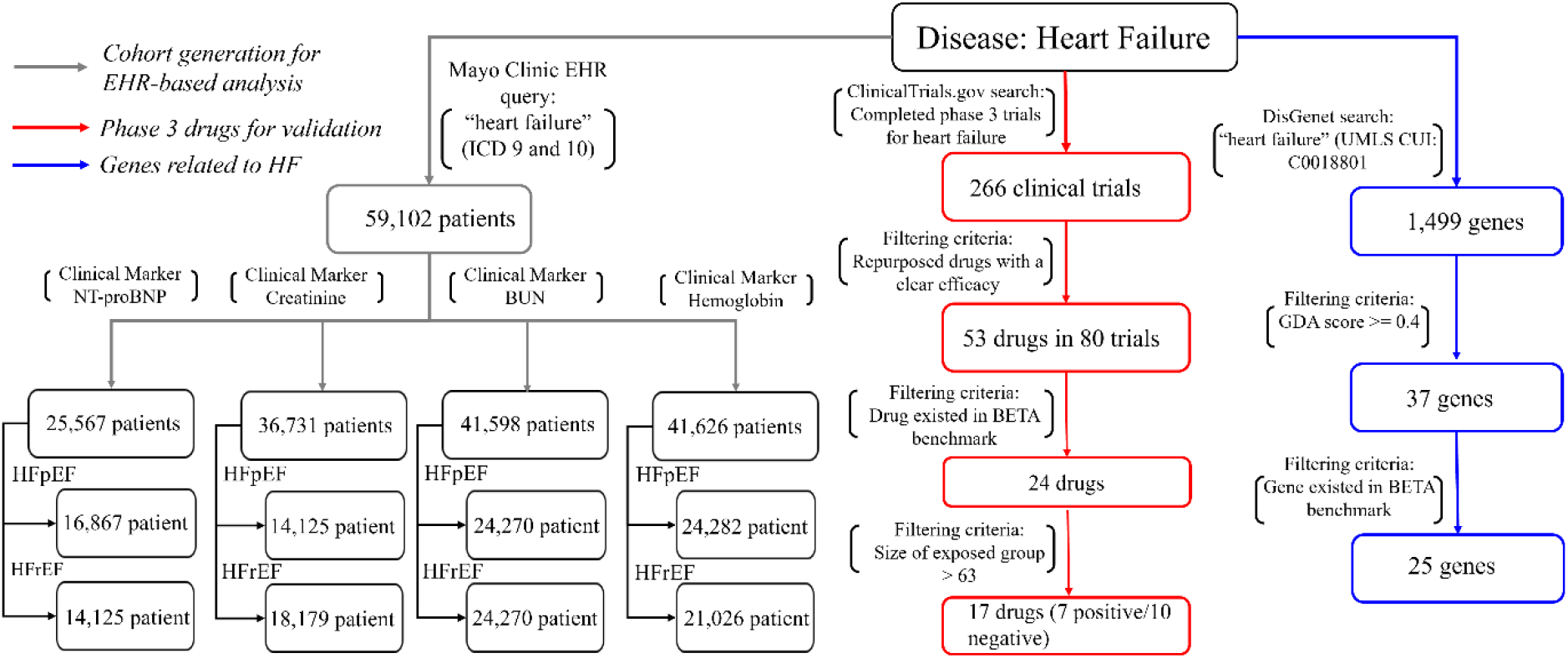
The flow of the data generation for the study.

## 3. Conclusion and Discussion

To bridge the gap between the outcomes of RCTs and those inferred from EHRs in the context of ET, this study introduces a new strategy for predicting the efficacy of repurposed drugs through the integration of biological and clinical insights. Our research addresses the previously unexplored accuracy of using EHRs for drug discovery in the adaptation of ETs^25, 26^. By evaluating 17 drugs repurposed for phase 3 HF RCTs, we highlighted the inherent limitations of trial simulations based solely on EHR data. We advocate for a more sophisticated approach that improves these simulations of RCTs with target prediction for repurposing HF drugs.

In the following sections, we will discuss several aspects and issues related to this study.

### AI-based RCT emulation

The gap between real-world evidence generation in clinical settings (e.g., EHR) and the controlled environment of RCTs means that EHR-based simulations cannot replicate RCT outcomes exactly^16, 17, 35^. Unlike existing studies^25, 26, 36^ that aimed to improve data generation through AI methods like the propensity score, our study sought to refine emulation through two primary strategies: 1) incorporating biological insights via target prediction, and 2) stratification based on variable interactions (e.g., HF medications and sex). Our approach demonstrated encouraging outcomes in predicting efficacy, surpassing baseline ET methodologies. While our experiment reported optimal results in **Section 2.4**, selecting the ideal combination of interactions is not straightforward. Therefore, we believe that the integration of KG in high-throughput drug screening could present a more viable solution. Furthermore, utilizing a more precisely tailored stratified cohort could be particularly advantageous for specific tasks in which the confounders and interacting variables within RCTs are well-defined. Developing a more sophisticated method to comprehend these variables is crucial for the process of data generation from EHR.

### Eligibility criteria for emulated trials

HF is a prominent health concern affected by a range of factors, including genetics, environmental influences, and various chronic and acute conditions^37, 38, 39, 40^. The drugs we evaluated from phase 3 trials were specifically chosen for their potential to address the underlying causes of HF and to alleviate symptoms, targeting different HF categories such as HFpEF, HFrEF, or HF in general. These trials often employed a variety of selection criteria, taking into account the study’s goals, potential risks, and confounding factors. In our research, we utilized demographic information, HF medications, HF subtypes, confounding variables, and prognostic markers to analyze drug efficacy through simulation. We recognize that this does not encompass all variables considered in the trials we reviewed. While including more variables in the simulations could reduce bias and improve accuracy, it might also diminish the generalizability, which is crucial for broadly screening drugs for clinical trials. Therefore, further research is necessary to determine the optimal way to select variables for simulated trials in the context of high-throughput drug screening.

### HF-related genes

The target prediction process is essential for evaluating the potential of repurposing drugs for new therapeutic uses. In prioritizing genes that might be targeted by drugs, our proposed method utilizes GDA scores^28^—derived from various scientific sources including the volume of supporting literature—as criteria for gene selection, treating all genes as equally significant in the prediction of drug-target relationships. Yet, the relevance of specific genes should be assessed based on their role in biological processes, such as those involved in pharmacokinetics and pharmacodynamics, to tailor treatments more closely to individual needs. For instance, previous research has indicated that genes like AGT and HIF1A are notably linked to an increased risk of HFpEF in individuals with chronic kidney disease^41^ and those suffering from obesity and metabolic syndrome^42^, respectively. Although the significance is based on the prediction of the potential novel drug-target associations, it may be beneficial to evaluate these genes more comprehensively as they are discovered to play a more critical role in drug development (e.g., AGT plays a central role in the regulation of blood pressure^43^, and HIF1A has been implicated in the development of cardiac hypertrophy and fibrosis at a low oxygen level^44^)

### EHR

Our study was conducted solely based on Mayo Clinic data, but there are more sophisticated approaches that can be explored by leveraging multiple EHR sources. There are two potential directions to consider. Firstly, utilizing integrated EHR databases, such as All of Us^45^, could offer a feasible solution to analyze a large patient cohort. However, this approach may introduce potential bias as the factors used to pull the cohort may vary across different participating hospitals. Secondly, sharing models, like federated learning^46, 47^, could be employed, where separate models are built with a selected set of hospitals within an existing research network, such as PCORnet^48^. However, this approach may require additional effort in data processing and could face common challenges, such as the competitive nature of maintaining advantages among participating sites^49, 50^. Furthermore, although a more advanced ad-hoc linear model could be utilized to select the combination of the cohort and potentially achieve better prediction performance, we did not include it in the results section due to concerns about overfitting and lack of generalizability. The focus of our study was to employ 17 drugs as a case study to evaluate the proposed method’s ability to predict the efficacy of the drugs for HF. While the initial results demonstrate promising predictive performance, drawing conclusions about the method’s effectiveness at this stage would be premature. Consequently, it is crucial to evaluate whether the proposed method can achieve similarly promising outcomes across a diverse array of diseases. This assessment will aid in refining the method’s effectiveness and promote its use in drug repurposing for precision medicine within clinical settings.

## 4. Methods

### 4.1. Repurposing drugs for HF in phase 3 RCTs

We conducted a search on *ClinicalTrials*.*gov* using the keywords “heart failure” and filtered by “completed” status and “phase 3” phase in order to acquire drugs undergone the phase 3 RCTs. Out of the total 266 clinical trials gathered, two experts, Drs. Chen and Dai, conducted reviews of each trial to assess whether it was specifically designed for HF and evaluated the repurposed efficacy. Their evaluations were based on the trial’s results or related articles associated with the clinical trial number. The reviews provided by both experts were subsequently merged for comparative analysis, and only those trials exhibiting consistent determinations were ultimately included. After this review, we narrowed down the list to 80 clinical trials for 53 drugs. Subsequently, an additional filtering process was conducted to exclude drugs that were not present in the BETA benchmark. This led to the identification of a list comprising 17 drugs. Out of the drugs we examined, 7 were determined to have beneficial impacts on HF, while the other 10 showed no discernible effects. For a comprehensive list of repurposed drugs, please refer to **Supplementary Table 1**.

### 4.2. Retrieve of the potentially druggable genes

Using the publicly available knowledgebase, DisGeNET^28^, we conducted a search for genes associated with HF. DisGeNET is a comprehensive database that contains genes and variants linked to human diseases, sourced from GWAS catalogs, scientific literature, and animal models. By entering the search term “heart failure” (UMLS CUI: C0018801) into the DisGeNET platform, we obtained a list of 1,499 results. We filtered them based on the GDA^28^ score, which is a confidence score calculated by the number and type of sources (level of curation, organisms). We identified 25 genes related to HF that were also present in the BETA benchmark. For more information on these genes, please refer to **Supplementary Table 2**.

### 4.3. Target prediction

Given a heterogeneous network *G*(*V, E*), a set of vertices *V* (i.e. biomedical entities), and a set of edges *E* (i.e. known associations), where *V* are multiple types of vertices (e.g., drugs or targets) and *E* are multiple types of edges that connect the vertices (e.g., drug-target associations), our objective is to predict the potential new associations among *V*. Specifically, we have four types of vertices, which are drugs *D*, targets *T*, diseases *Ds*, and side effects *S*, where *D* ∈ *V, T* ∈ *V, Ds* ∈ *V, S* ∈ *V*. For each drug *d*, a chemical structure of such a drug is given as *str*_*d*_. For each target *t*, a protein sequence of such a target is given as *seq*_*t*_. When there is a linking edge existing in the *G*, the pair of vertices is defined as {(*u, v*)|*u* ∈ *V* ∧ *v* ∈ *V* ∧ (*d, t*) ∈ *E*}, while there is an unknown linking edge, the pair of vertices is defined as {(*u, v*)|*u* ∈ *V* ∧ *v* ∈ *V* ∧ (*d, t*) ∉ *E*}. Therefore, the problem of drug-target prediction can be defined as, given a pair of a drug *d* and a target *t*, predicting whether the pair {(*d, t*)|*d* ∈ *V* ∧ *t* ∈ *V*} is an existent association (referred to as positive) or nonexistent association (referred to as negative). We have developed a novel hybrid model for drug-target prediction with the incorporation of two encoding networks (as depicted in **Figure 1(b)**), the D-network that learns the biological features from drug chemical structure and protein sequence, the N-network that learns the topological features from a set of bipartite knowledge graphs. Specifically, for the D-network, we get the latent vectors (i.e., embeddings) generated from the encoder *ℛ*_*drug*_(*str*_*d*_) for the drug’s chemical structure. Similarly, the latent vectors for protein sequences are obtained from the encoder *ℛ*_*target*_(*seq*_*t*_). In practice, we adopted the encoder layers from DeepPurpose^32^. For the N-network, we get the embedding of a node *v* from the t-th layer as *f*^*t*^(*v*) = ∑_*e*(*v,v*′)∈*E*_ *f*^*t*−1^(*v*^′^), *f*^*t*−1^(*v*^′^) embeddings of the neighborhood *v*^′^. In practice, *f*^*t*^(*v*) is implemented based on NeoDTI^31^. We further updated the embedding as *f*^t^(*ν*) ← *Concat*(*f*^*t*−1^(*ν*), *ℛ*), where *ℛ* is an embedding learned from D-network. Specifically, *ℛ* = *ℛ*_*drug*_ when *ν* is a drug and *ℛ* = *ℛ*_*target*_ when *ν* is a target. The output of the designed network is 𝕔 which corresponds to the set of bipartite graphs. The loss function is 𝕔 = λ_1_𝕔_1_ + λ_2_𝕔_*q*_ … + λ_*k*_𝕔_*k*_, where λ_1_ + λ_2_ + ⋯ + λ_k_ = 1. 𝕔_*k*_ the objective function for the subgraph *k*, which is calculated: 𝕔_*k*_ = ∑_*e*(*u,v*) ∈*E*_‖*s*(*e*) − f(*u*)ℚℕ^*T*^f(*v*)‖^2^, where *s*(*e*) is the weight of an edge *e* and *s*(*e*) = 1 if there is a link between the nodes *u* and *v*. f(*u*) is the embedding of node *u*, ℚ and ℕ are the edge projection function. For a detailed description of the proposed model, please refer to **Supplementary File 1**. For the evaluation of target prediction, please refer to Section 2.2.

### 4.4. Efficacy analysis based on EHR

We conducted a comparative analysis, utilizing methods such as the Chi-square test, to assess whether the cohort exposed to the repurposed drug under investigation would achieve more favorable outcomes compared to the unexposed cohort, which consists of patients not receiving the repurposed drug. The EHR data of Mayo Clinic HF patients utilized in this study were obtained with the appropriate approval from the Mayo Clinic’s Institutional Review Board (IRB). We used four specific tests that are considered as the HF prognostic markers in clinical^37, 40, 51^: N-terminal pro-b-type natriuretic peptide (NT-proBNP), blood urea nitrogen (BUN), creatinine, and hemoglobin. The values of the clinical results were categorized as normal or abnormal based on normal ranges defined in clinical practice. The HF patients are defined by ICD9 and 10 codes (see **Supplementary Table 3** for the details), and the following four medications recommended by the AHA/ACC/HFSA guideline^40^ were considered as the baseline HF drugs (see **Supplementary Table 4** for the details) for the unexposed cohorts: ACEI, BB, ARB, and LD. Our exposed group consisted of HF patients who also take repurposed drugs before completing their medication regimen from any of the above categories. We stratified the patients based on the clinical subtypes of HF, which are HFrEF and HFpEF. Instead of using a static subtype for all drugs across the evaluations, we also incorporated a dynamically stratified patient group called the BASELINE group, which was defined based on the eligibility criteria specified in RCTs. (see **Supplementary Table 1** for the BASSELINE cohort for each drug). In addition to the three biological variables, which are age, sex, and ethnicity, we also include 12 confounding factors that are defined by our domain experts (Drs. Li, Dai, Chen, and Bielinski) in this study. Please refer to **Table 1** for the cohort details.

Given that the advanced method to compute the propensity score did not yield improved results in balancing variables^26^, we adopted the traditional logistic regression-based propensity score model to balance the confounders before the analysis. Specifically, propensity score is generated based on the logistic regression in prediction of the drugs, which is further used to generate the pair-wised case/control groups for analysis^52^. Subsequently, we analyzed the exposed and unexposed groups to learn the difference in the distribution of normal versus abnormal values for prognostic markers (such as NT-proBNP) through chi-square testing, where a value of 1 indicates a significant positive correlation, -1 denotes a significant negative correlation, and 0 signifies a lack of significant correlation (refer to Sections 2.2 and 3.3 for results). To expand our analysis and incorporate the effects of interactions between HF medications (namely ACEI, BB, ARB, and LD) and biological factors (such as sex), we employed a binomial regression model to detect the positive or negative impacts of the drugs under investigation. The final prediction value will be an integration of the coefficients derived from the binomial regression model into the target prediction based on 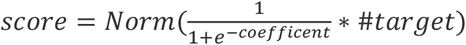, where *coefficent* is the coefficient from the binomial regression model and #*target* is the number of positive targets predicted and *Norm*(∗) is a normalized score.

To calculate AUCROC and PRAUC, we compared the normalized prediction scores from the proposed model and the chi-square test with the labeled drugs against the labeled drugs (i.e., a positive effect and no effect).

## Data Availability

Obliged by the Mayo Clinical data sharing policy, we cannot share the patient data.

## Contributors

NZ conceptualized the study. NZ designed the target predictive model, SZ and SC developed and optimized the model. SC prepared EHR data and NZ conducted the experiments. NZ, SC, and YY conducted the statistical analysis and data visualization. The initial draft of the report was composed by NZ and SC, and the other authors evaluated and provided feedback on it. All of the authors collectively assumed the ultimate responsibility of deciding to submit the report for publication and committed to being answerable for all aspects of the work, ensuring that any concerns regarding the precision or honesty of any part of the work are adequately addressed and resolved. NZ supervised and administered the study.

## Acknowledgment

This work is supported by a grant from the National Institute of Health (NIH) NIGMS (R00GM135488).

## Data and Code Availability

Data used in the preparation of this article were obtained from the BETA, which can be accessed in https://github.com/bioIKEA/IKEA_BETA_Benchmark. The used code is publicly available at https://github.com/bioIKEA/BETA_Trial_Prediction

